# SCLERODERMA AND MALIGNANCY

**DOI:** 10.1101/2021.01.25.21250479

**Authors:** Jaime A. Vondenberg, David R. Cisneros, Adnan N. Kiani, Maheswari Muruganandam, Sharon E. Nunez, N. Suzanne Emil, Wilmer L. Sibbitt

## Abstract

**Background/Objectives:** Describe clinical characteristics of malignancy-associated scleroderma in a regional population.

**Methods:** Scleroderma patients with current or past malignancy were prospectively identified from a cohort of 125 scleroderma subjects of a largely Hispanic population. Demographic information included scleroderma subtype, serologies, cancer diagnosis, age at diagnosis, co-morbid conditions, therapies, and survival outcomes.

**Results:** 16.0% (20/125) of scleroderma patients were identified as having a malignancy. The mean age of malignancy onset and scleroderma onset were 55.6±10.3 and 52.7±12.9 years respectively (95% CI of difference: -4.6 <2.9< 10.4, p=0.44). Antinuclear antibody was positive in 70%, anticentromere antibody in 35.0%, and anti-topoisomerase antibodies in 25%. Tumor proportions were adenocarcinomas 55.0% (11/20) (1 gastric, 2 pancreatic, 1 lung, 2 thyroid, 5 breast), 15% (3/20) squamous cell carcinoma (1 skin, 1 uterine cervical, and 1 rectal), and 20% (4/20) benign tumors (2 meningiomas, 1 renal adenoma, 1 plasma cell). Age of onset of scleroderma greater than 42 years respectively accounted for 85% of all cancer cases. The standardized incidence ratio (SIR) was 2.0 (CI at 95%: 1.2<2<3.1), indicating 100% increase in cancer risk. Treatment of the malignancy did not resolve the scleroderma in any of the cases. Five-year survival after cancer diagnosis was 70.0%, although 2 of the survivors had tumor recurrence.

**Conclusions:** Malignancy, especially adenocarcinoma, occurs frequently in scleroderma in minority populations with up to 16% of patients affected. High-risk patients are those with scleroderma-onset greater than 42 years in whom routine screening for common cancers should be undertaken.

## Background

Scleroderma is an aggressive immune-mediated disease characterized by progressive fibrosis of the skin and visceral organs [1]. Compared to the general population, multiple studies have shown that scleroderma patients are at an increased risk of cancer [2-8]. Epidemiologic studies have demonstrated a unique temporal clustering of the onset of scleroderma to the development of cancer, suggesting that underlying malignancy might be instigating the onset of scleroderma or scleroderma and the medications used to treat scleroderma have been inciting malignancy [4-24]. In patients with concurrent malignancy and scleroderma, both conditions may improve with cancer therapy further suggesting that scleroderma, in certain cases, may be a true paraneoplastic syndrome [4]. Recent tumor marker and modeling studies have demonstrated that malignancies may be present subclinically years prior to malignancy diagnosis [25-27]. Thus, cancer associated with scleroderma may be more common than previously recognized [4, 25-28]. We report a new series of malignancy associated with scleroderma in a minority population comprised predominately of Hispanics and Native Americans in the Southwest United States.

## Methods

This study was approved by the institutional review board (IRB) – the Human Research Review Committee (HRRC) of the University of New Mexico Health Sciences Center and all subjects provided informed written consent before all procedures and examinations. The study adhered to the principles in the Declaration of Helenski. 125 scleroderma patients were clinically examined and prospectively identified by one attending rheumatologist during normal clinical care and diagnosed as having scleroderma using the 2013 Classification Criteria for Systemic Sclerosis [29]. Patient demographics are shown in Table 1. All patients underwent a standard clinical autoimmune diagnostic laboratory panel consisting of the following autoantibodies: 1) antinuclear antibody (ANA) including titer and pattern, 2) anti-DNA antibody, 3) anti-Smith antibody (Sm), 4) anti-RNP antibody, 5) anti-centromere antibody, 6) anti-Jo1 antibody, 7) anti-scleroderma (anti-topoisomerase) antibody, 8) rheumatoid factor, and 9) anti-CCP antibody. The inclusion criteria were as follows: 1) the subject at least 18 years old, 2) diagnosis of scleroderma, and 3) a malignancy diagnosis. Exclusion criteria included patients less than 18 years, pregnant females, prisoners, and anyone who could not give consent. Twenty total patients with both malignancy and scleroderma diagnoses were prospectively identified and collated during normal clinical care from the total clinical cohort of 125 scleroderma patients. From these cases further data was retrospectively collected regarding the following: age at scleroderma diagnosis, age at cancer diagnosis, time between the diagnosis of scleroderma and cancer, classification of scleroderma, histologic type of cancer, co-morbid conditions, therapies used for treatment of scleroderma, treatment used for malignancy, and positive serologies.

**Table 1.** Patient Demographics

|  |  |
| --- | --- |
| Patients with cancer | 16% (20/125) |
| Mean observation time | 12.5±7.9 years |
| Mean age of scleroderma diagnosis | 52.7±12.9 years |
| Mean age of cancer diagnosis | 55.6±10.3 years |
| Mean age at last visit | 62.1±9.3 years |
| Women | 90% (18/20) |
| Men | 10% (2/10) |
| Hispanics | 65% (13) |
| Native American | 10% (2) |
| Non-Hispanic Caucasian | 20% (4) |
| Asian | 5% (1) |
| Diffuse Scleroderma | 65% (13/20%), |
| Limited Scleroderma | 35% (7/20) |
| Adenocarcinoma | 55.0% (11/20) |
| Squamous cell carcinoma | 15% (3/20) |
| Benign tumors | 20% (4/20) |
| Survival at 5 years after cancer diagnosis | 70.0% (14/20) |

### Statistical Methods

Standard summary statistics (means, proportions) were calculated and statistical analysis including standardized incidence ratio (SIR) was performed with Simple Interactive Statistical Analysis (SISA) (Consultancy for Research and Statistics, Lieven de Keylaan 7, 1222 LC Hilversum, The Netherlands; http://www.quantitativeskills.com/sisa/) using control malignancy data from New Mexico from the American Cancer Society [30]. Statistical differences between measurement data were determined with Student t-test. P values <0.05 were considered significant.

## Results

The demographics of the patient population is shown in Table 1. Mean observation time of the cohort was 12.5±7.9 years. 16.0% (20/125) cases of the total 125 scleroderma population were diagnosed with concomitant malignancy indicating a malignancy rate of 100 new malignancies/100,000 individuals/year compared to the greater New Mexico malignancy rate which is 49 new malignancies/100,000/year with a standardized incidence ratio (SIR) of 2.0 (CI at 95%: 1.2<2<3.1), indicating an excess of 100% in new cancers in the scleroderma cohort using 2018 statistics [29]. Thus, this case series of malignancy-associated scleroderma included 20 patients: 10 with diffuse scleroderma, 9 with limited scleroderma, and 1 with overlap scleroderma. 18/20 (90%) were women and 2/10 were men (10%). Of the 20 patients, 13 (65%) were Hispanic, 1 (5%) was Asian, 4 (20%) were White non-Hispanic Caucasians, and 2 were Native Americans (10%) reflecting local care population. The mean age of the patients at last clinical visit was 62.1±9.3 years. The mean age at diagnosis of malignancy was 55.6±10.3 years old and the mean age at diagnosis of scleroderma was 52.7±12.9 years old (95% CI of difference: -4.6 <2.9< 10.4, p=0.44); thus, the ages of onset on scleroderma and malignancy were very similar across the population and shown in Figure 1. Patients with onset of scleroderma at greater than 37 and 42 years old respectively accounted for 95% and 85% of all cancer cases.

**Figure 1.**
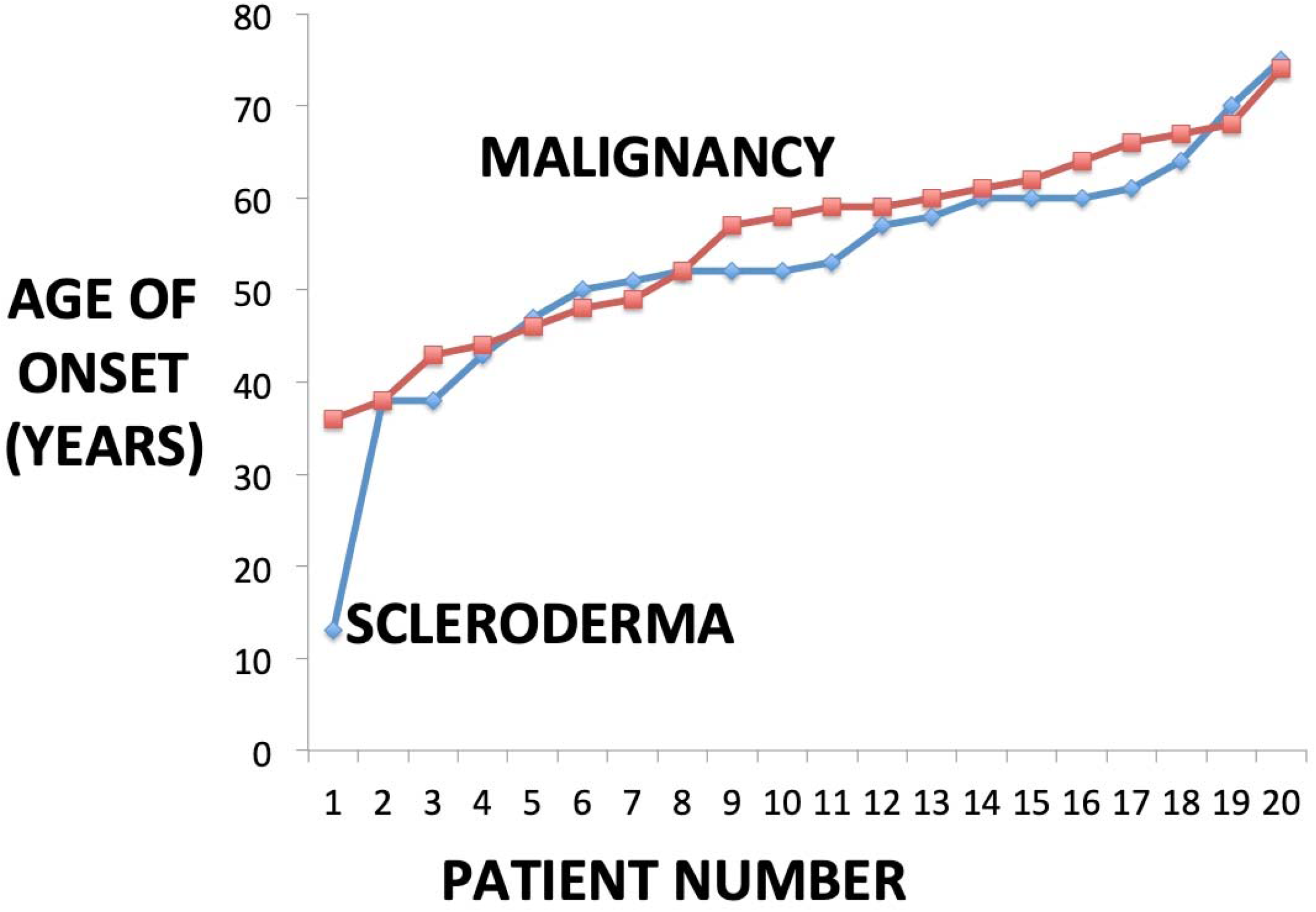
Age of Onset of Malignancy and Scleroderma. The graph shows the entire 20 subject scleroderma and malignancy cohort and demonstrates that the ranked age of onset of scleroderma (mean: 55.6±10.3 years old) and the ranked age of onset of malignancy (mean: 55.6±10.3 years old) curves are very similar across the population (95% CI of difference: -4.6 <2.9< 10.4, p=0.44)

Common scleroderma manifestations associated with malignancy included distal scleroderma 20/20 (100%) (Figure 2), proximal scleroderma 17/20 (85%), cutaneous telangiectases 17/20 (85%) (Figure 3), gastroesophageal reflux disease (GERD) 20/20 (100%), Raynaud’s phenomenon 19/20 (95%), interstitial lung disease (pulmonary fibrosis) 11/20 (55%), and pulmonary artery hypertension 8/20 (40%). Mean estimated pulmonary artery pressure by transthoracic echocardiogram at last observation was 32.6±16.6 mm Hg.

**Figure 2.**
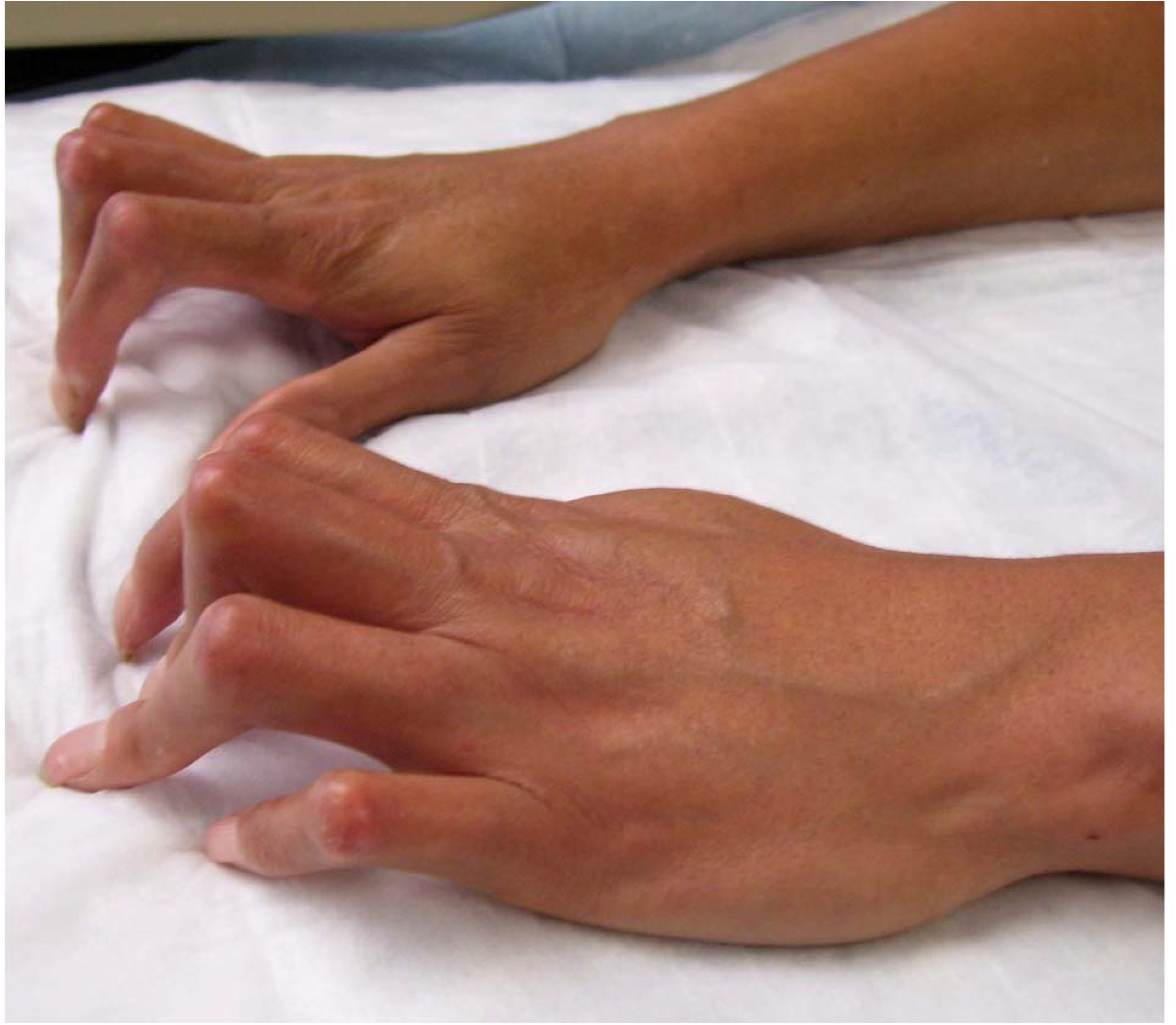
Distal Scleroderma with Cancer. The photograph demonstrates distal sclerodermatous changes in the hands with contractions of the small joints of the fingers in a patient with thyroid adenocarcinoma.

**Figure 3.**
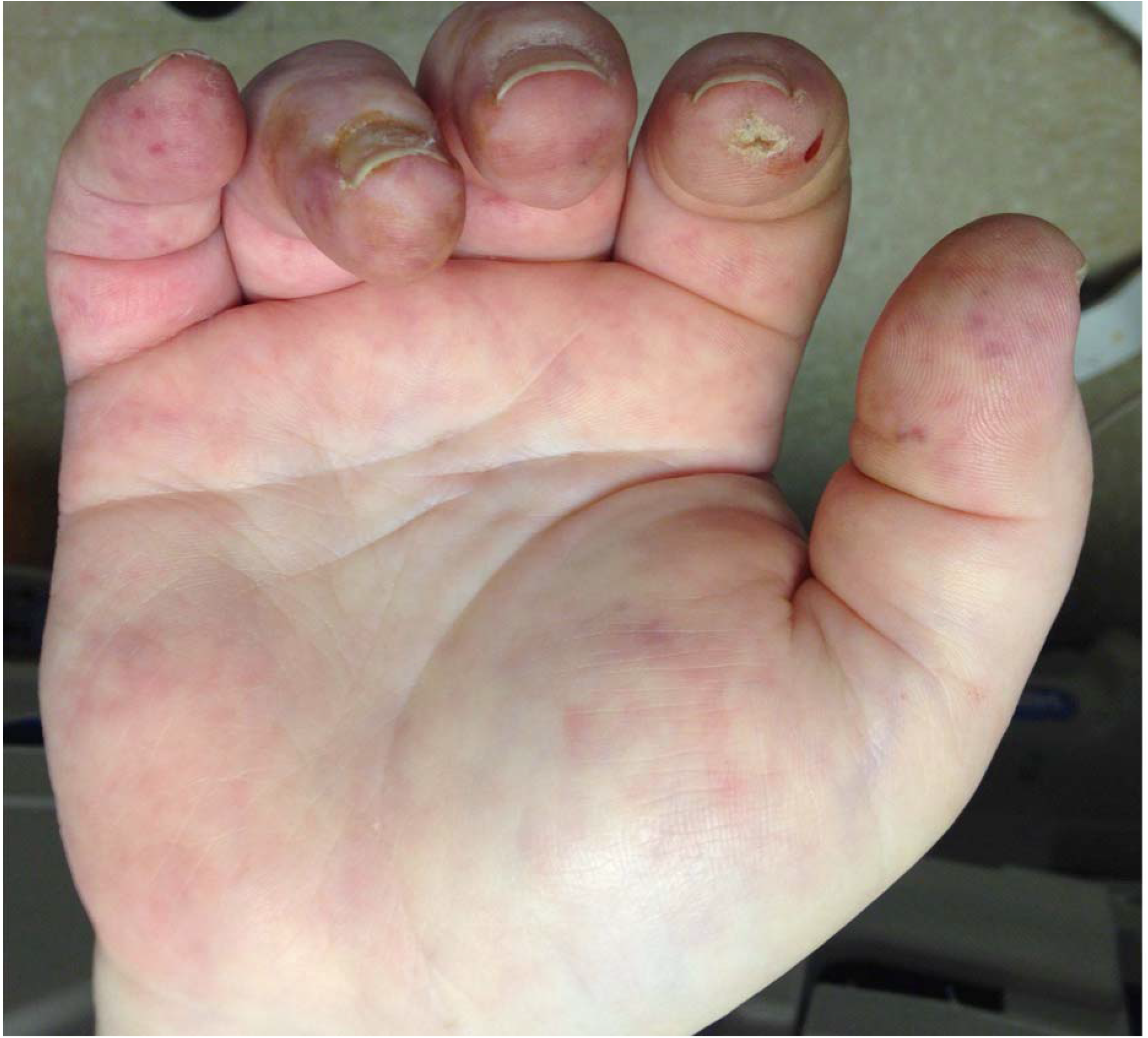
Malignancy-Associated Scleroderma with Telangiectases. This palmar view demonstrates scleroderma of the hand, distal ulcerations on the finger pad from vasospasm, contractures and multiple telangiectases in a patient with thyroid adenocarcinoma.

**Figure 4.**
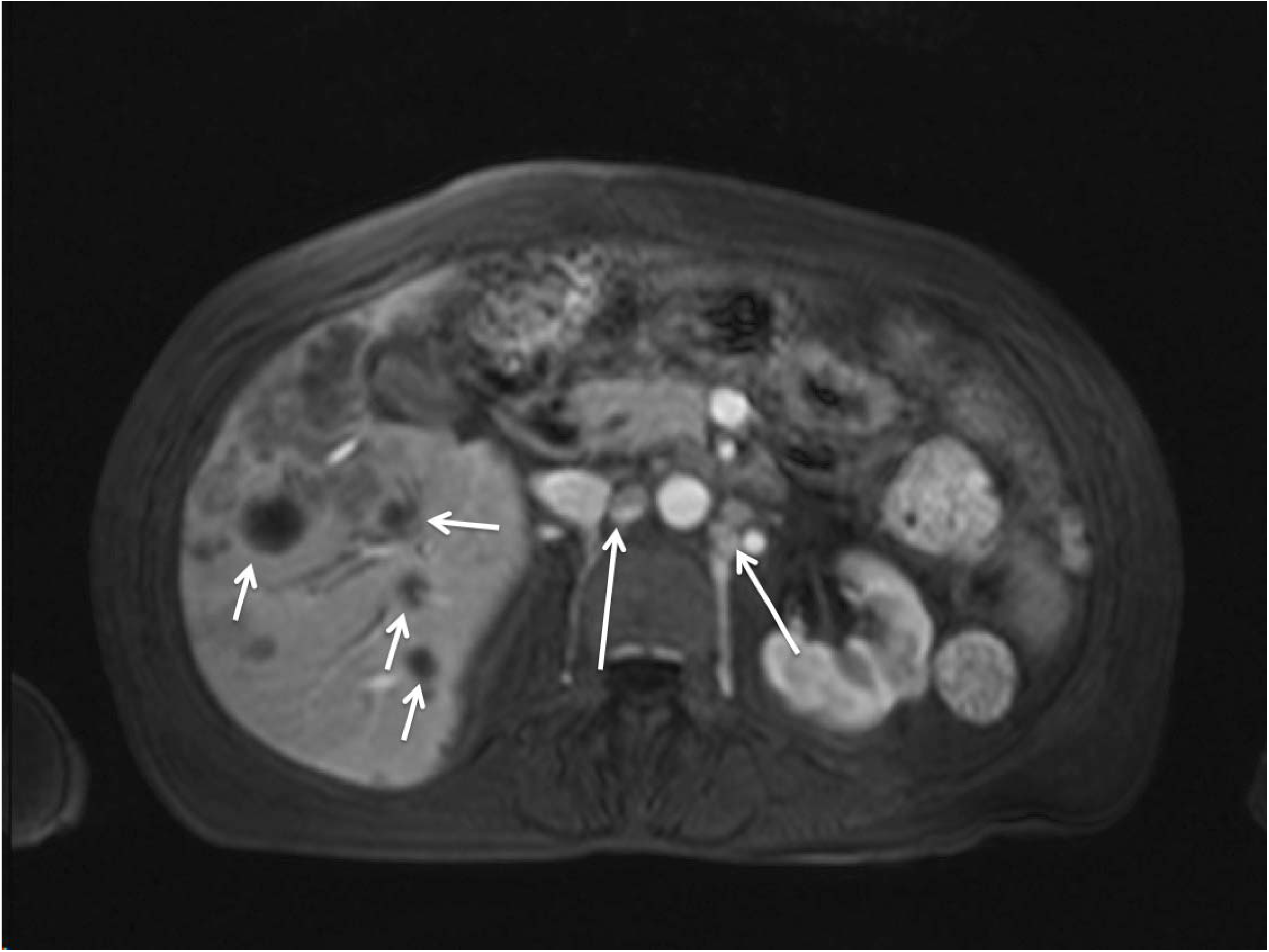
Malignancy-Associated Scleroderma with Multiple Metastases. This computed tomographic image demonstrates a scleroderma patient suffering with metastatic pancreatic adenocarcinoma in the periaortic lymph nodes (long arrows) and multiple metastases in the liver (short arrows).

Serologic testing was typical for scleroderma. Antinuclear antibody (ANA) was positive 14/20 (70%), with a mean inverse titer of 275.1±235. ANA patterns included: speckled 3/14 (21.4%), homogeneous 2/14 (14.3%), centromere 7/20 (35%), and nucleolar 1/14 (7.1%). Positive specific serologies in this cohort also included: anticentromere antibody 7/20 (35%), anti-scleroderma (anti-topoisomerase) antibody 5/20 (25%), rheumatoid factor 2/20 (10%), anti-Jo1 antibody 0/20 (0%), anti-DNA antibody 1/20 (5%), anti-Smith antibody (Sm) 1/20 (5%), anti-RNP antibody 1/20 (5%), and anti-CCP antibody 0% (0/20).

A wide variety of tumor types were observed. 55.0% (11/20) of tumors were adenocarcinomas (1 gastric, 2 pancreatic, 1 lung, 2 thyroid, 5 breast), 15% (3/20) squamous cell carcinoma (1 skin, 1 uterine cervical, and 1 rectal), and 20% (4/20) were “benign tumors” (2 meningiomas, 1 renal adenoma, 1 plasma cell). Thus, scleroderma was associated with both malignant neoplasms as well as “benign” neoplasms.

7/20 (35%) of patients were diagnosed with cancer prior to scleroderma (mean 5.2 ± 6.8 years prior to scleroderma diagnosis). 3/20 (15%) were diagnosed with cancer simultaneous to the scleroderma, and 10/20 (50%) were diagnosed with cancer after the scleroderma diagnosis (mean 9.1±6.8 years after the scleroderma diagnosis). Certain therapies for cancer have been shown to induce a scleroderma-like state and interestingly, the nine patients diagnosed with cancer prior to scleroderma had received some combination of surgery, radioactive iodine, external radiation, tamoxifen, or anastrozole [22, 31-35]. The treatment of malignancy did not lead to the resolution of the scleroderma in any of the subjects (i.e., all 20 patients continued to have scleroderma after cancer therapy). After 5 years of the diagnosis of cancer, 14/20 (70.0%) of affected patients were still alive, though 2 had had tumor recurrence. Six patients (30%) died within 5 years of the tumor diagnosis: 1 from pulmonary hypertension, 1 from congestive heart failure, and 4 from metastatic cancer. 12/20 (60 %) scleroderma-malignancy cases remain tumor-free to date.

## Discussion

The present case series demonstrates 20/125 (16.0%) of scleroderma patients in a distinct local population with a predominance of individuals of Hispanic and Native American descent developed a malignancy in average observation of time of 12.5 years. It should be noted that the local Hispanic population typically also has Native American ancestry ranging from 15-80% [36]. This malignancy prevalence is similar to other reports cancer in scleroderma ranging from 7.1-20.0% [1-8, 15-24]. Malignancy-associated scleroderma is defined by a cancer diagnosis that precedes, is simultaneous with, or appears after the diagnosis of scleroderma and suggests that malignancy can be an important risk factor, association, or marker for the emergence of scleroderma and vice-versa (Figure 1) [4, 26-28].

In our series, 70% of malignancy-associated scleroderma patients were ANA positive, 35.0% were anticentromere antibody positive, 25.0% were anti-topoisomerase antibody positive, and 30.0% were seronegative which is similar to prior studies [1-7]. In the present series, 50% (10/20) of patients had diffuse scleroderma, 45% (9/20) had limited scleroderma, and 5% (1/20) had an overlap scleroderma similar to other scleroderma-cancer cohorts [16-24]. Compared to the general population, several studies have shown that scleroderma patients are at an increased risk of concomitant cancer, the timing of which can be variable [4, 11-23]. Bonifazi et al found that the relative risk of developing all invasive cancers in scleroderma was 1.75 with the strongest association with lung cancer (RR 4.35) but also showed a significant increase in risk with hematologic malignancies (RR 2.24) [14]. Pooled data from multiple studies have shown significant increases in the risk of cancer of the lung, liver, hematologic system, and bladder, as well as of non-Hodgkin’s lymphoma and leukemia [1-7,13-24]. Our cohort demonstrated tumor proportions with only 5% developing lung cancer and 5% with hematologic malignancies while adenocarcinomas dominated perhaps reflecting the different genetics or environment of our study population.

The risk of malignancy has been reported to be more pronounced in patients with diffuse cutaneous disease, and in patients with positive RNA polymerase III antibodies, especially within a few years of scleroderma onset [13-24, 38]. Our study demonstrated that the mean onset of scleroderma (52.7±12.9 years) and the diagnosis of malignancy (55.6±10.3 years, p=0.47) were very similar, supporting this type of close relationship as shown graphically in Figure 1.

Additional factors contributing to the development of malignancy in scleroderma patients could relate to the chronic inflammation caused by the scleroderma, the cytotoxic medications used in the management of scleroderma or a genetic susceptibility that might induce both cancer and scleroderma [1-23]. Cyclophosphamide and mycophenolate are frequently used in the treatment of scleroderma, and cyclophosphamide exposure has been associated with both hematologic and bladder malignancies [9, 10] whereas mycophenolate has been tied to non-melanoma skin cancer and lymphoma [11-13]. In contemporary practice, scleroderma is most commonly treated with mycophenolate while use of cyclophosphamide has markedly declined over the last decade, thus none of our patients received cyclophosphamide but many received mycophenolate.

The inflammation associated with cancer or the cancer therapy itself might damage the endothelium and cause vascular disease and thus elicit Raynaud’s phenomenon and fibrosis comparable to that seen in scleroderma; this would be supported by the population of patients who develop scleroderma after their cancer diagnosis [4, 9-12, 30-39]. Drugs that have been associated with cutaneous, vascular, and pulmonary complications resembling scleroderma include bleomycin, docetaxel, paclitaxel, carboplatin, gemcitabine, uracil-tegafur, radiotherapy, and graft versus host disease after bone marrow transplantation [4, 30, 31, 39].

More recently, immune checkpoint inhibitors including nivolumab, pembrolizumab, and others have also been associated with the development of scleroderma and scleroderma-like skin changes during cancer therapy [33,34]. Some of the patients in this case series received docetaxel, paclitaxel, carboplatin, or radiotherapy for their cancer. Of those patients who developed scleroderma during or after cancer therapy, drug- or radiation-induced scleroderma cannot be entirely excluded, although in all subjects scleroderma remained after conclusion of cancer therapy suggesting that the cases were predominately primary scleroderma.

In certain patients with synchronous malignancy and scleroderma diagnosis, scleroderma can improve with treatment of the cancer suggesting that scleroderma in these cases may be a true paraneoplastic syndrome [4,15,16, 24,34,35]. The fact that in the present series the scleroderma persisted after successful cancer treatment and that most patients have classic scleroderma subtypes suggests that in this series the subjects had true scleroderma as opposed to a reversible scleroderma-like paraneoplastic syndrome.

Adenocarcinomas of various origins occurred in 11/20 patients (55%) of this series, and thus adenocarcinomas dominated the present cancer-scleroderma association. The dominance of adenocarcinomas of this series diverges from prior reports in that only (5%) had lung carcinoma associated with scleroderma [8]. The patient population of this case series included predominantly Native American and Hispanics with considerable Native American ancestry of the Southwest United States, an understudied population in terms of rheumatic diseases including scleroderma, but also a population with lower rates of tobacco use [36]. A wide variety of tumor types were observed including both malignant neoplasms and benign neoplasms similar to previous larger series [2-24]. Benign neoplasms including renal adenoma had been reported previously, but unlike the present report few previous reports included meningioma as a scleroderma-associated tumor; yet in the current series 10% were meningiomas [2-24].

A significant proportion of the patients (60%) in this case series developed cancer prior to or simultaneous with the scleroderma diagnosis. This is consistent with recent advances in the understanding of long latency of malignancy before diagnosis, where the cancer cells are present latently 1.5 to 10 years before the neoplasm is clinically diagnosed [4, 25-27]. Several meta-analyses have found that the risk of cancer diagnosis *following* scleroderma diagnosis was greatest within the first 12 months and up to 2 years [4, 15]. Our series is similar in that 50% of the patients who developed cancer after scleroderma diagnosis did so within the two-year window.

The causation of scleroderma is unknown; however, overproduction of collagen is thought to result from an autoimmune dysfunction, in which the immune system starts to attack the kinetochore of the chromosomes [1,34,35,37,38]. A significant player in the process is transforming growth factor (TGFβ) which is overproduced, and the collagen-secreting fibroblast overexpresses the receptor for TGFβ [38]. Apart from TGFβ, connective tissue growth factor (CTGF) also has an important role [37]. Indeed, a particular *CTGF* gene polymorphism is present at an increased rate in systemic sclerosis [38]. An intracellular pathway (consisting of *SMAD2/SMAD3, SMAD4* and the inhibitor *SMAD7*) is in part responsible for the secondary messenger system that induces transcription of the proteins and enzymes responsible for collagen deposition.

Jimenez and Derk proposed three main theories of scleroderma causation: 1) the fibrotic process is induced by a physical agent, and all other changes are secondary or reactive to this direct physical insult, 2) phenotypic alterations in genetically susceptible cells effectuate DNA changes which alter the cell’s behavior inducing the fibrotic process and 3) fetomaternal cell transfer causes microchimerism, with a second trigger leading to a type of graft-versus-host reaction that causes the actual development of the disease [37].

Recent gene profiling has demonstrated that scleroderma patients often express known oncogenic gene signatures suggesting that the susceptibility genes for scleroderma and the susceptible genes for cancer may overlap [35]. Dolcino and colleagues found up regulation of genes involved in apoptosis, including upregulation of B-cell lymphoma 2 (BCL2); BCL2-like 1 (BCL2L1); BCL2-like 13 (BCL2L13); mitochondrial fission factor (MFF); translocase of inner mitochondrial membrane 50 homolog (TIMM50) (involved in breast cancer), and zinc finger and BTB domain containing 16 (ZBTB16) [35]. These researchers also found overexpression of genes involved in cell proliferation including MYC proto-oncogene, bHLH transcription factor (MYC); v-akt murine thymoma viral oncogene homolog 2 (AKT2); Pim-1 proto-oncogene, serine/threonine kinase (PIM); cyclin D3 (CCND3), and T-cell leukemia/lymphoma 1 A (TCL1A). Similarly, Joseph and colleagues found genetic alterations of the polymerase III polypeptide A (POLR3A) locus in peripheral blood lymphocytes of scleroderma patients with antibodies to RNA polymerase III who developed cancer but not in scleroderma patients without antibodies to RNA polymerase III who did not develop a malignancy, suggesting that POLR3A mutations may be associated with the development of cancer in scleroderma patients [23].

As reviewed by Bianchi et al and Jimenez et al one suspected mechanism behind the autoimmune phenomenon that induces scleroderma is the existence of microchimerism, where fetal cells circulate in maternal blood, triggering an immune reaction to what is perceived as foreign material [37, 40]. An analogous clinical microchimerism is graft-versus-host disease after bone marrow or other tissue transplant that can result in scleroderma-like changes with fibrosis of the skin and internal organs [39]. Recent tumor marker and modeling studies have demonstrated that malignancies are typically present subclinically up to 10 years before diagnosis; thus, cancer co-emerging with scleroderma may occur more often than has been previously recognized [4, 25-27]. Indeed, circulating malignant cells are often identified in blood before the clinical diagnosis of cancer or scleroderma, suggesting that microchimerism where the circulating cells are malignant or premalignant and induce similar reactive immune changes may also in part explain the relationship between scleroderma and malignancy [28]. These observations support the hypothesis of a common primary event early in oncogenesis that could make a self-antigen become immunogenic, and next trigger an autoimmune response against both tumor cells and normal host tissues inducing generalized tumor-associated scleroderma [23,34].

Thus, the observation of cancer-scleroderma concurrence may reflect not only the known carcinogenic effect of chronic inflammation, but also true activation of oncogenes and circulating malignant, pre-malignant, and autoreactive cells creating microchimerism that are important for both the autoimmune scleroderma inception and also for the initiation and proliferation of malignancy [23,34,35,40]. The present study shows a close concurrence between mean scleroderma onset (52.7±12.9 years) and mean malignancy onset (55.6±10.3 years, p =0.47) supporting these hypotheses as shown in Figure 1.

There are a number of shortcomings to this report. First, this is a cross-sectional study from rheumatology patient panel of 125 scleroderma patients; not a true regional study of scleroderma, bringing up the possibility of selection bias. Certainly, the study needs to be expanded to determine whether the entire local scleroderma population in the region reflects a similar increased malignancy rate and we are planning to pursue this. Secondly, although a complete clinical autoantibody panel including anti-centromere antibodies and anti-topoisomerase antibodies were obtained in all cases, RNA III polymerase antibodies were not a component of this standard panel and thus were not obtained in all patients, but have recently been included in the diagnostic panel [5].

Strengths of the this report include 1) a unique series with a predominance of previously underreported Hispanic and Native American subjects with malignancy-associated scleroderma, 2) the relative dominance of adenocarcinomas and breast cancer and relative lack of lung cancer as in many previous reports, 3) the association of meningiomas with scleroderma, and 4) the high prevalence of cancer diagnosis in the local cohort scleroderma population at 16% which should increase with time as the cohort is followed.

## Conclusions

Scleroderma often occurs before, simultaneously with, or after the development of malignancy. In this underreported predominately Hispanic and Native American scleroderma population, the prevalence of malignancy is quite high at 16% indicating that cancer screening with the diagnosis of scleroderma should be considered in these populations. In general, the observed cancers are common malignancies that standard breast, thyroid, skin, lung, and colorectal cancer screenings may detect. Further, the high resolution computed tomography (CT) of the thorax that is typically performed to evaluate for the presence and extent of interstitial lung disease may also provide an additional layer of cancer screening. This study also suggests that if intractable or atypical headaches are present in scleroderma patients, meningiomas should also be excluded by appropriate imaging.

## Key Points

1.) Malignancy frequently occurs prior to, simultaneously with, and after the diagnosis of scleroderma. Clinicians should be aware of the scleroderma-malignancy association to allow for focused appropriate screening for early diagnosis and therapy of both disease processes.

2.) Scleroderma onset 42 years and older appear to be at particular risk for malignancy, and these high-risk populations should undergo appropriate age-related screening.

3) When CT scanning is obtained to assess the extent of interstitial lung disease in a scleroderma patient, findings of nodules, irregularities, or lesions in solid organs should prompt further investigation.

## Data Availability

Deindentified data are available as requested as permitted by the local IRB - the Human Research Review Committee (HRRC) of the University of New Mexico Health Sciences Center.

